# qMRI-BIDS: an extension to the brain imaging data structure for quantitative magnetic resonance imaging data

**DOI:** 10.1101/2021.10.22.21265382

**Authors:** Agah Karakuzu, Stefan Appelhoff, Tibor Auer, Mathieu Boudreau, Franklin Feingold, Ali R. Khan, Alberto Lazari, Christopher J. Markiewicz, Martijn J. Mulder, Christophe Phillips, Taylor Salo, Nikola Stikov, Kirstie Whitaker, Gilles de Hollander

**Affiliations:** NeuroPoly Lab, Institute of Biomedical Engineering, Polytechnique Montreal, Montréal, QC, Canada; Montreal Heart Institute, Montreal, QC, Canada; Center for Adaptive Rationality, Max Planck Institute for Human Development, Berlin, Germany; NeuroModulation Lab, School of Psychology, University of Surrey, Guildford, UK; Stanford University, Stanford, CA, USA; Department of Medical Biophysics, Robarts Research Institute, University of Western Ontario, London, Canada; Wellcome Centre for Integrative Neuroimaging, FMRIB, Nuffield Department of Clinical Neurosciences, University of Oxford; Department of Experimental Psychology, Utrecht University, Utrecht, the Netherlands; GIGA Cyclotron Research Centre in vivo imaging, GIGA Institute, University of Liège, Liège, Belgium; Florida International University, Miami, FL, USA; The Alan Turing Institute, London, UK; Zurich Center for Neuroeconomics (ZNE), Department of Economics, University of Zurich, Zurich, Switzerland; Spinoza Centre for Neuroimaging, Amsterdam, The Netherlands

## Abstract

The Brain Imaging Data Structure (BIDS) established community consensus on the organization of data and metadata for several neuroimaging modalities. Traditionally, BIDS had a strong focus on functional magnetic resonance imaging (MRI) datasets and lacked guidance on how to store *multimodal* structural MRI datasets. Here, we present and describe the BIDS Extension Proposal 001 (BEP001), which adds a range of quantitative MRI (qMRI) applications to the BIDS application sphere. In general, the aim of qMRI is to characterize brain microstructure by quantifying the physical MR parameters of the tissue via computational, biophysical models. By proposing this new standard, we envision standardization of qMRI which makes multicenter dissemination of interoperable data possible. As a result, BIDS can act as a catalyst of convergence between qMRI methods development and application-driven neuroimaging studies that can help develop quantitative biomarkers for neural tissue characterization. Finally, our BIDS extension offers a common ground for developers to exchange novel imaging data and tools, reducing the practical barriers to standardization that is currently lacking in the field of neuroimaging.

## Introduction

The brain imaging data standard (BIDS) is an open source initiative from the neuroimaging community that aids in standardizing neuroimaging data sets. BIDS was originally developed with functional MRI (fMRI) applications in mind, describing experimental task blocks in relation to a hierarchical organization of reconstructed MR images^1^. This convention engaged researchers to share hundreds of open fMRI data on the openneuro platform^2,3^ and develop interoperable processing workflows that can seamlessly process these datasets^4^. Popular examples include the MRIQC^5^ and fmriprep^6^ pipelines, which can be executed even online for any valid BIDS fMRI dataset. Similarly, the development of an MRI k-space data standard, ISMRM-RD^7^, led open-source MRI reconstruction packages to adapt this convention and now aids potential users in performing advanced reconstruction tasks with minimal effort ^8,9^. These success stories from open science exemplify how data standards can change the landscape of community-driven software for the better, leading to a collective change in researchers’ behaviour to adhere with FAIR (findability, accessibility, interoperability and reusability) principles of scientific data^10^. Here we present our work extending the BIDS to include multi-contrast MRI acquisitions. BIDS Extension Proposal 001 (BEP001) was merged into the standard (on 23 February 2021) and focuses on quantitative MRI (qMRI) applications.

Quantitative MRI methods map physical magnetic properties of the (brain) tissue. Their application consists of two steps: i) collecting multiple MRI images, where the contributions of effective micrometer-level MRI parameters is systematically manipulated by adapting very specific acquisition parameters, and ii) fitting the resultant voxel intensity variations across the images to a computational (biophysical) model^11^. The results are a single or multiple quantitative *map of the estimated parameters* across the imaged volume. The effective MRI parameters that are typically studied include longitudinal and transverse relaxation time constants (T1 and T2, respectively), proton density (PD), magnetization transfer (MT), and local diffusion coefficient (e.g., fractional anisotropy, FA, or mean diffusivity, MD). The multi-parametric mapping^12^ (MPM) protocol offers a set of acquisitions that can quantify more than one MR parameter at a time. Another popular technique used in qMRI is field mapping, which characterizes inhomogeneities in MRI radiofrequency (RF) transmit (B1+) and receive (B1-) profiles, as well as static magnetic field (B0) to correct qMRI parameter estimation errors due to these field inhomogeneities.

The earliest qMRI applications date back to the late 70’s^13^ and primarily focused on relaxometry, mapping of quantities such as T1 and T2* relaxation time. Since then, the field has witnessed multiple waves of methods development, driven by technological advances and emerging trends in MRI research^14,15^. Recently, with the surge of deep learning methods, the gamut of parameter estimation methods have become much larger than ever before ^15-19^. Interestingly, however, we still do not precisely know the healthy range of relaxation time values in a multi-center setting^20^ nor do we know how to establish diagnostically reliable tissue typing protocols. This discrepancy highlights that multicenter standardization should be a critical step toward evaluating the clinical potential of decades-long improvements in the acquisition and processing of qMRI data.

Under more controlled research settings, qMRI offers obvious advantages over conventional MRI contrasts (e.g. T1 weighted images) in structural feature extraction. Given that MRI is not a direct measurement of in vivo anatomical structures, voxel-wise morphometry analyses are subjected to various biochemical and physiological confounders affecting the voxel intensity^21^.Hence, the capacity of disentangling MRI signal components lands qMRI as a more reliable approach to study structural variations^22^. This makes qMRI particularly useful for comparisons of the brain anatomy of different (clinical) groups^23-25^ and for more consistent, unbiased automated anatomical segmentation^26-29^. The same principle can be exploited to make qMRI sensitive to tissue microstructure, such as iron concentration or myelination. Recent meta analyses revealed that a majority of qMRI methods are comparably sensitive to the myelin content^30,31^, although certain parameters such as myelin water fraction (MWF, relaxometry-based) and macromolecular pool fraction (MPF, MT-based) appear to be more specific.

Given the advantages offered by parametric maps in providing structural information and the current landscape of myelin imaging methods, it seems likely that many more myelin imaging methods leveraging the potential of qMRI will be developed in the future. This leads to one of our four main motivations behind covering qMRI methods in BIDS: to bring FAIR principles to a variety of qMRI data that are finding widespread use in neuroimaging research. Other motivations include i) driving open-source qMRI tools to adapt a consolidated input/output convention, ii) creating standardized databases that can help simplify the use of qMRI in clinical and translational research, and iii) stimulating an open provision of qMRI data that can be collected by imaging equipment that is available to a small group of researchers.

Drawing upon the principles outlined in BIDS, we introduce the first consensus data and metadata organization standard for qMRI. This work is a culmination of years of effort (the earliest drafts of the BEP-001 date back to 2017) and discussion between neuroimaging researchers and MRI methods developers around the globe. Our extension will not only aid in organizing qMRI data, but will also facilitate multi-center collaborative work, encourage neuroscientists to adapt advanced MR techniques and go a long way toward the standardization of qMRI methods.

## Results

### A new BIDS common principle: entity-linked file collections

The majority of qMRI methods necessitate the grouping of a set of similar images where specific acquisition parameters are carefully varied. Furthermore, the images that are collected for qMRI application do not usually have a clear “weighting” description (e.g., T1w, T2w), like conventional structural images. The novel concept of file collections decouples the semantics of logical group identification from contrast weighting labels or acquisition sequence names that are not originally developed for qMRI (e.g., FLASH). Instead, suffixes for such logical units may indicate a generic MRI readout type (e.g., multi-echo gradient echo: MEGRE), a qMRI sequence name (e.g, magnetization prepared two rapid gradient echoes, MP2RAGE) or a qMRI data collection framework (e.g., variable flip angle, VFA). Table-1 lists file collection suffixes for various qMRI and fieldmap data, and the quantitative parameters they can derive. These suffixes span a wide range of qMRI applications including relaxometry, MT imaging, multiparametric mapping, and RF field mapping. Application scope can be extended without necessarily adding more suffixes. The BIDS qMRI appendix presents a set of rules and suggestions to add new qMRI suffixes to the specification (https://bids-specification.readthedocs.io/en/stable/99-appendices/11-qmri.html).

**Table 1-.**
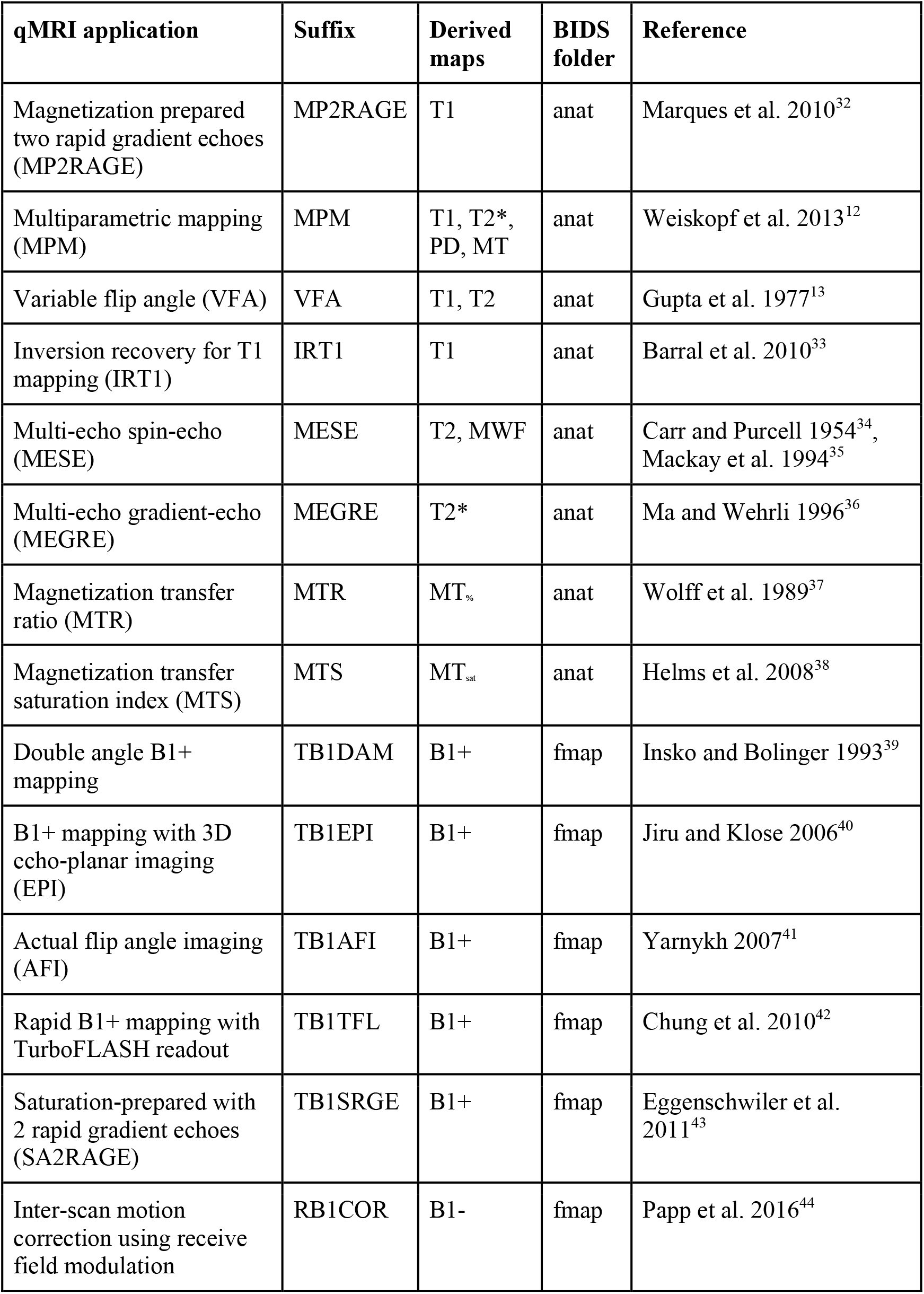
File collections of anatomy imaging data to derive parametric maps of longitudinal, transverse and observed-transverse relaxation times (T1, T2 and T2*, respectively), proton density (PD), magnetization transfer ratio and saturation index (MTR and MTsat) and myelin water fraction (MWF). Relaxation rates (e.g., T1^-1^ and T2^-1^) and residual terms (e.g., M0) are excluded from the table for brevity.

Note that the use of file collections is not exclusive to qMRI, anatomy imaging data, or even MRI. Any imaging modality calling for a file grouping logic to define a quantitative or qualitative application can benefit from this principle by specifying a descriptive suffix and filename entity. Such changes would require additional BIDS extensions to create a valid file collection.

To distinguish individual files of a file collection, we introduced filename entities that are associated with commonly altered acquisition parameters (e.g., flip angle) or with inherent components of the same data (e.g., phase information), hence the name “entity-linked file collection” (Table-2).

**Table-2.** Filename entities representing an MRI acquisition parameter or designating an inherent part of the reconstructed image (e.g., magnitude or phase).

| Entity format | Entity values | Associated acquisition parameter | Associated qMRI file collections |
| --- | --- | --- | --- |
| echo-<index> | 01,02,03,...,n | EchoTime | MEGRE, MESE, MPM |
| flip-<index> | 01,02,03,...,n | FlipAngle | VFA, MTS, MPM |
| inv-<index> | 01,02,03,...,n | InversionTime | IRT1, MP2RAGE |
| mt-<label> | on/off | MTState | MTR, MTS, MPM |
| part-<label> | mag/phase | N/A | MP2RAGE |

It is important to highlight that these entities cannot store acquisition parameter values in the filename, but can only index or categorize them. Respective parameter values are stored in so-called “sidecar JSON”-files. Requirement level of these entities in relation to file collections are presented in the BIDS entity table appendix (https://bids-specification.readthedocs.io/en/stable/99-appendices/04-entity-table.html).

### Data organization for qMRI file collections and quantitative parametric maps

By combining entities in the filename that represent different acquisition parameters (Table-2) with entity-linked file collection suffixes (Table-1), BEP001 provides an intuitive way to organize filenames of most existing qMRI data. For example, raw data from MP2RAGE acquisitions comprises both magnitude and phase reconstructed images, acquired at two successive inversion times (Fig-1a). The respective file collection for MP2RAGE (Fig-1c) clearly defines these components via *part* and *inv*-components, which are required for the MP2RAGE file collection. Note how the BIDS inheritance-rules do allow for using a single JSON-file to describe both phase- and magnitude-images, since these have identical acquisition parameters. In addition, the same collection suffix can be extended to specify its multi-echo variant^45^ using the echo entity, which is made optional to MP2RAGE. For clarity, these specific use cases are defined in the BIDS qMRI appendix.

**Figure-1.**
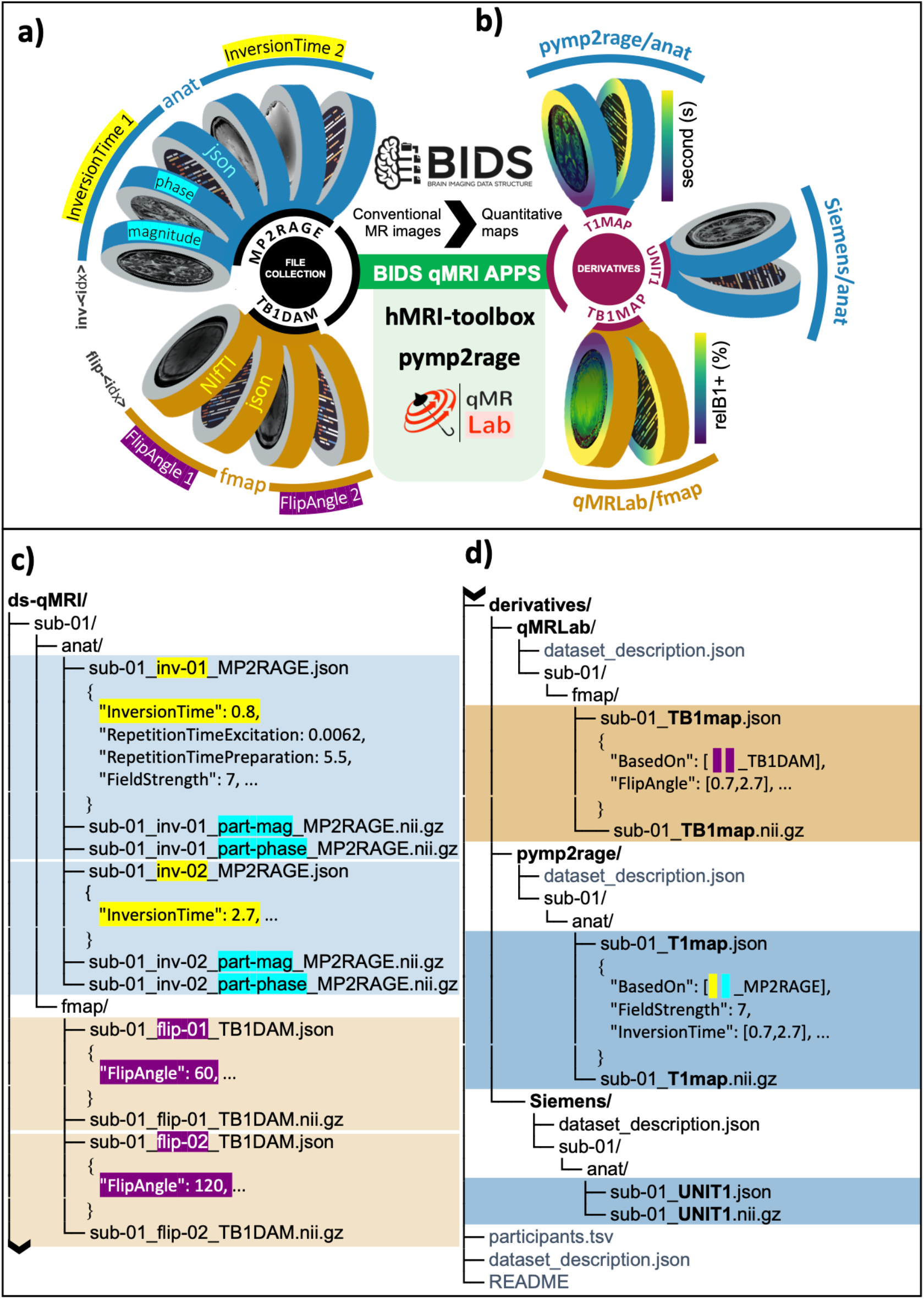
**a)** Schematic of BIDS formatted raw quantitative MRI (qMRI) data representing MP2RAGE (anat) and TB1DAM (fmap) file collections, for which entity-linked metadata fields are highlighted for the InversionTime (yellow), the FlipAngle (purple) and for the reconstructed image type (cyan). **b)** Derivatives of MP2RAGE and TB1DAM file collections generated by using pymp2rage and qMRLab to calculate T1 and B1+ maps, respectively, including a vendor-native derivative of UNIT1 images. **c)** File organization of raw qMRI data for MP2RAGE and TB1DAM file collections, where respective linking entities are highlighted for the inv entity (yellow, InversionTime), the flip entity (purple, FlipAngle) and the part entity (cyan, magnitude/phase). **d)** File organization of qMRI derivatives indicating how sidecar JSON files of quantitative maps generated by open-source software keeps a log of the input files (the BasedOn field) and associated acquisition parameters (FlipAngle in TB1map and InversionTime in B1map).

The same logic applies to the raw images of double-angle B1+ mapping, identified by the TB1DAM suffix (Fig-1c). In this case, the maximum value of the flip entity indicates that the data is collected over two flip angles. We recognize that an alternative approach to organize such data is stacking images at each flip angle into the 4th dimension of a Nifti-file, and storing the corresponding metadata in vector form using a single JSONfile. This approach offers a less crowded file list for this particular example. However, indexing acquisition parameter dependent variations across additional dimensions is less favorable for comprehensive qMRI methods. For example, MPM collects raw data at different echo times, flip angles and MT preparations with the option of phase reconstruction. After extended debates that took more than a year, the qMRI-BIDS extension group ultimately concluded that this approach is less favourable for human-readability of qMRI datasets, especially for multiparametric acquisition methods where the number of images per protocol can go into the dozens.

### Metadata requirements for file collections and quantitative parametric maps

For the file collections, linking entities (Table-2) indicate a requirement for the respective acquisition parameters that are subject to change from image-to-image. Therefore, the entity table appendix lists such parameters as required in relation to the corresponding file collection suffix based on the descriptions made in the BIDS schema. Note that not all the parameters that change across file collection images are captured by a linking entity, but may still be required for data fitting. For example, the value of the *FlipAngle* parameter might (but does not necessarily) covary with that of *InversionTime* between MP2RAGE file pairs; however, the filenames are distinguished solely by the *inv* entity (since that is the crucial parameter that is swept over, whereas the flip angle could in principle remain the same). In addition, certain parameters that are constant across file collection images may be required as well. For example, *RepetitionTimeExcitation* and *RepetitionTimePreparation* are required metadata for an MP2RAGE acquisition. Such parameters are required when they are strictly necessary to calculate the qMRI-maps that a specific acquisition scheme was designed to obtain; e.g., a T1-map in case of MP2RAGE. BEP001 added an array of new metadata fields that may be required for certain file collections (e.g. *MTState*, specifying whether an MT preparation is enabled in an MPM acquisition, associated with the *mt* linking entity) or provide supporting information (e.g., *SpoilingRFPhaseIncrement*, specifying the amount of incrementation applied to the phase of an excitation pulse). The complete list of metadata fields and their requirement levels for all the qMRI file-collections are included in the BIDS release v1.5.0 and later. Currently, metadata conversions for some of these required fields have been implemented in dcm2niix^46^, a commonly used DICOM to NIfTI converter to create BIDS-compatible datasets.

Certain quantitative parameters cannot be interpreted in absence of fundamental scanner specifications. For example, to interpret relaxometry maps (e.g., T1map), the magnetic field strength must be known. The BEP001 ensures that such requirements are met (again, see the qMRI Appendix in BIDS release v1.5.0 and later). Moreover, sidecar JSON files of quantitative maps contain all the metadata values involved in the fitting by representing varying parameters in vector form and inheriting the constant ones from the raw images. To supplement the provenance recording of parameter estimation process with software-relevant details, the derived dataset and pipeline rules are respected as outlined in the modality agnostic files section of the main specification.

Finally, the units and range of the fitted parameters have been standardized by BEP001 to define interchangeable qMRI maps. For relaxometry-based parameters (e.g., T1map or T2map), the time is described in seconds and the rate in reciprocal seconds or Hz. Wherever applicable, unitless ratio maps are described in percentage (e.g., MTRmap or MWFmap). For quantitative susceptibility maps (i.e., Chimap) the local magnetic susceptibility is represented in parts per million. The RF transmit maps (i.e., TB1map) are specified in relative percentage units, where 100% denotes the ideal case (i.e., measured flip angle equals the nominal value). Any deviations from 100% convey proportional deviations from the intended field strength. Please note that certain quantitative parameters are described in arbitrary units, where the acceptable range of values vary based on the target anatomy (e.g., MTsat).

### Community software and the role of BIDS in standardizing qMRI

As of release v1.5.0, the BIDS validator can perform on BEP001-compatible qMRI data at the directory and filename level rules, based on the entity requirement levels specified per file collection suffix. However, metadata-level validation rules have not been implemented yet. This is mainly because multi-vendor extraction of qMRI related metadata fields (e.g., MTState or RepetitionTimePreparation) is not supported by commonly used converters. Recently, we started working with dcm2niix^46^ and BIDSme (https://github.com/CyclotronResearchCentre/bidsme) developers to identify and map vendor-specific header information to BEP001-compatible metadata.

Nevertheless, some metadata entities that are of profound importance to the accuracy of quantitative maps cannot be typically found in the vendor-native DICOM headers. For example, the BIDS fields of *RFSpoilingPhaseIncrement* and *SpoilingGradientMoment* are two major determinants of T1 and B1+ estimation accuracy using spoiled gradient echo based applications^47^. Although this information is not provided by vendors, open-source pulse sequence development frameworks such as Pulseq^48^, PyPulseq^49^, Gammastar^50^, TOPPE^51^, SequenceTree^52^, ODIN^53^ and RTHawk^54^ can make a qMRI-tailored metadata annotation possible. An example implementation is qMRPullseq, a collection of publicly available vendor-neutral pulse sequences that are designed to export images in accordance with BEP001 format without hidden acquisition parameters^55^. We highly encourage open-source MRI pulse sequence developers to use and contribute to the qMRI metadata annotations. This simple consensus can remove proprietary roadblocks from disseminating qMRI datasets that incorporate key information on the reproducibility of data acquisition.

Most qMRI methods can benefit from a plethora of BIDS applications^4^ to prepare data for parameter estimation and downstream statistical analyses. There are several open-source tools emerging to perform qMRI fitting at multiple levels, like the hMRI-toolbox^56^, qMRLab^55^, QUIT^57^, PyQMRI^58^, QMRTools^59^, mrQ^60^, Madym^61^, MITK-ModelFit ^62^, ROCKETSHIP^63^, DCEMRI.jl^64^ and DCE@urLAB^65^. Giving these tools the ability to operate on BIDS formatted data is an important step towards establishing interoperable qMRI processing pipelines.

## Conclusion

Quantitative MRI offers a rapidly developing set of techniques that can inform us about brain (micro)structure beyond what conventional MRI techniques have to offer^66^. We believe that, in coming years, qMRI will become increasingly important to both clinical and fundamental brain science. Therefore, a concrete standard for organizing and thereby also disseminating open qMRI data sets is much warranted. BEP001 extends the framework of the existing and very successful BIDS standard, to develop a standard for qMRI in the form of a “BIDS extension proposal”. To aid actual user adoption of this standard, it includes very precise descriptions of how to use it in many real-life qMRI use-cases, as well as many example data sets.

Currently, obtaining qMRI data is still expensive and needs considerable expertise, which is not readily available at many MRI facilities. Therefore, we also hope that BEP001 will aid researchers that do not have easy access to such facilities to get familiar with qMRI data and potentially can even use open qMRI data sets for their particular research questions.

Finally, the popularity of BIDS is likely in large part also due to some software packages that are designed around this standard and therefore extremely easy-to-use, when one’s data adheres to the BIDS standard^67^. We hope that the success of BIDS in the domain of functional MRI will also inspire and encourage MRI software developers to work on similar “BIDS apps” to make it easier to work with qMRI data, as well as make processing pipelines more open and transparent.

## Data Availability

During and since the development of BIDS extension proposal 001, multiple data sets have been converted to the new qMRI standard, in part also to stress-test the developing file naming schemes. Table-3 shows an (non-exhaustive) list of currently available qMRI data sets that are converted to the extended BIDS standard (release 1.5.0).

**Table-3.**
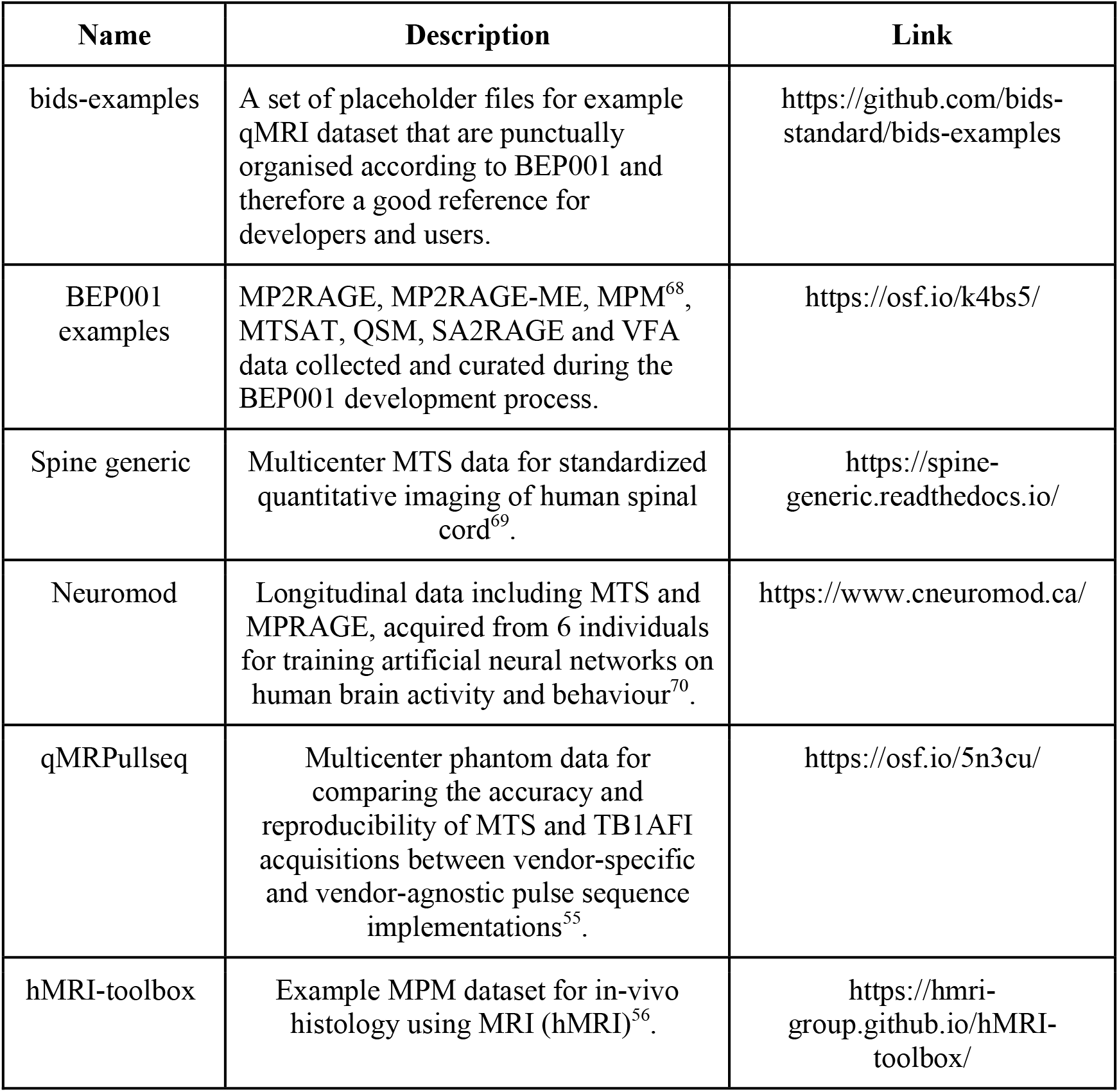
Various resources for example BIDS datasets making use of the specifications introduced by the BEP001 extension proposal.

## Methods

### Community-driven development of BEP001

The development history of BEP001 spanned nearly 5 years. This extension was initiated following mailing list discussions about standardizing MP2RAGE^32^ datasets and including multi-echo MRI acquisitions in late 2016 (https://bit.ly/bids_mailing). These discussions revealed that BIDS was still lacking a generic convention for specifying structural acquisitions yielding multiple contrasts. In the summer of 2018, meeting were held to hear concerns and questions from interested participants and to set an action plan for the development during the annual INCF NeuroInformatics conference in Montréal/Canada (http://www.neuroinformatics2018.org/) and the OHBM meeting in Singapore (https://www.humanbrainmapping.org/i4a/pages/index.cfm?pageID=3821). As a first action, a joint-community meeting was organized between MRI and neuroimaging scientists on 4 October 2018 (https://www.ismrm.org/virtual-meetings/virtual-meetings-archive/), where a consensus decision was made on extending the specification for a variety of qMRI methods. After this meeting, a standard operational procedure was established and followed to advance the proposal, focusing on both transparency and accessibility to other researchers (Fig. 2).

**Figure-2.**
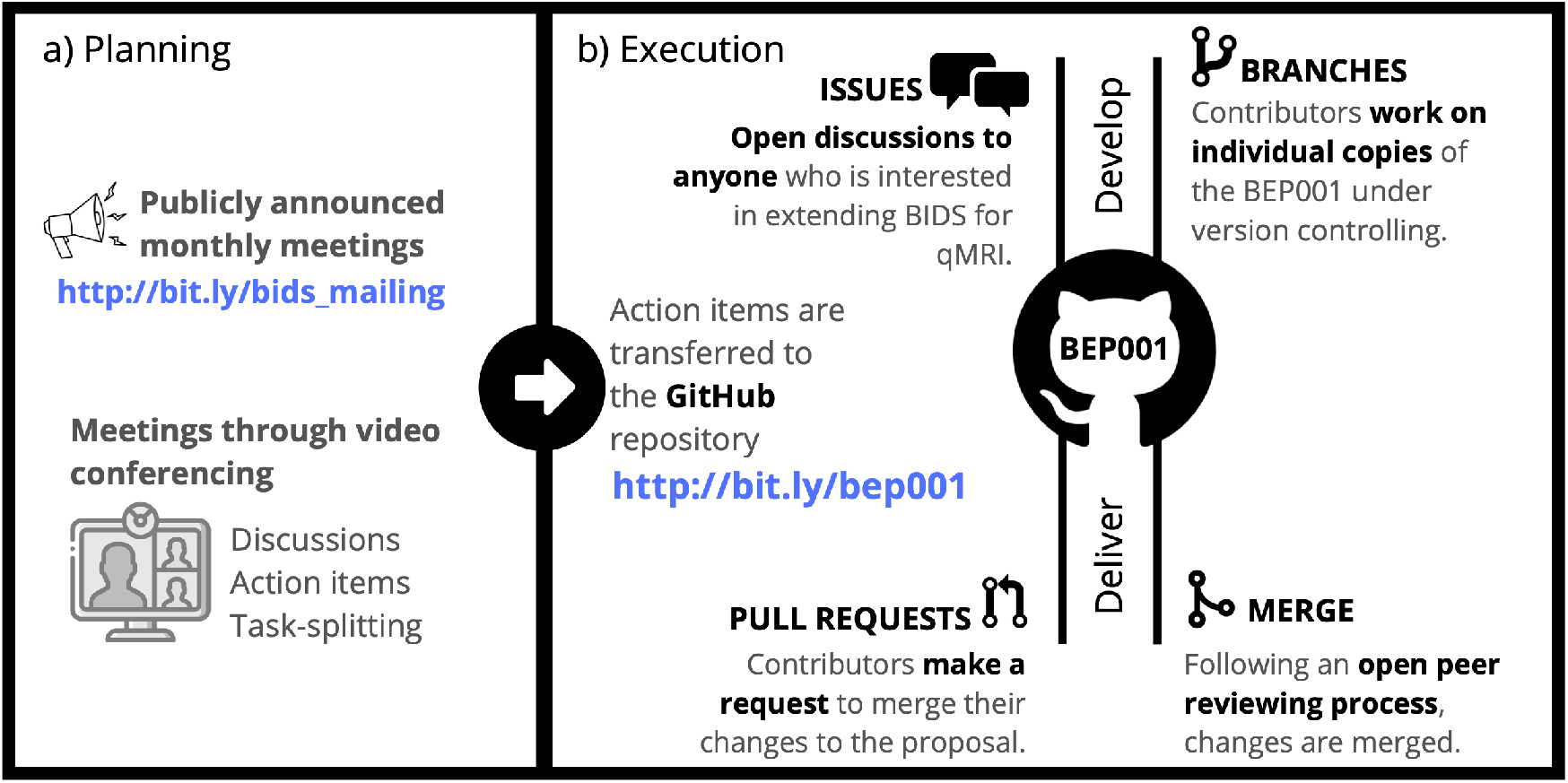
Summary of the standard operational procedure for improving BEP001. Outcomes from the monthly meetings **(a)** are transferred to a central GitHub repository, opened for more elaborate public discussions via issues and merged into the proposal through peer-reviewed pull requests **(b)**. BEP001 is inclusive to all communities who would like to contribute to the proposal or keep themselves up-to-date with the latest developments.

Interim outcomes from the development were presented in the 2020 annual conferences of OHBM^71^ and ISMRM^72^ to reach out more neuroimaging and MRI physics researchers, respectively. Following another year of development on the specification, example datasets and applications, BIDS incorporated and released BEP001 as part of their version 1.5.0. The main problems identified and resolved during the development are outlined in the following section, laying out the methodology of how qMRI can be incorporated into BIDS.

### Extending an existing standard for new use cases

BIDS traditionally focused on conventional anatomical images that are collected in functional MRI experiments and whose contrast characteristics are well-defined (i.e., mostly T1-weighted images). This posed a challenge for the naming scheme of collections of multimodal images used in qMRI. Unlike conventional structural data, qMRI inputs are usually formed by collections of images where specific acquisition parameters are systematically manipulated. Moreover, the line separating contrast characteristics between these images is blurred. A concrete example: in a multi-echo GRE acquisition with a long TRs, early echoes will be mostly PD- and B1+/B1-signal-weighted, whereas later echoes will be increasingly T2*-weighted. Most echoes will show a contrast that is the result of a mixture of underlying physical properties. This ambiguity disqualifies MRI weightings (e.g., T1w or T2starw) as suffix labels to specify interchangeable qMRI datasets. The use of often proprietary acquisition sequence names like “FLASH’’ (fast low angle shot) or “GRE” (gradient-recalled echo) as a suffix turned out to also be undesirable, because different MRI vendors use different naming conventions and, moreover, one type of sequence can often be used for numerous qMRI applications. To address this problem, BEP001 introduced a new common principle: file collections.

A second challenge that BEP001 addressed pertains to standardizing the data organisation of quantitative parametric maps. One central challenge of such maps is that the calculations on which they are based can be made both by proprietary vendor software run on the scanner system, or offline using open-source workflows. The resultant map can be described as derivative data in either case, yet the former lacks provenance of the whole calculation process and may not export the raw inputs to the calculation.

## Data Availability

https://github.com/bids-standard/bids-examples

https://doi.org/10.17605/OSF.IO/K4BS5

## Acknowledgements

The authors would like to acknowledge the work by other contributors to BIDS, and in particular those that contributed to BEP-001 via the Github repository, intermediate meetings, as well as a first draft on Google Drive. For BEP-001, recorded contributions include those from Suyash Bhogawar, Julien Cohen-Adad, Elizabeth Dupre, Chris Gorgolewski, Daniel Handwerker, Michael Harms, Ilana Leppert, Tobias Leutritz, Dylan Nielson, Julien Sein, Isla Staden, Wietske van der Zwaag, and Tobias Wood.

This research was funded in part by the Wellcome Trust [Grant 109062/Z/15/Z to AL]. For the purpose of Open Access, the author has applied a CC BY public copyright licence to any Author Accepted Manuscript version arising from this submission.

TA’s work has been funded by the Biotechnology and Biological Sciences Research Council, London (BB/S008314/1).

C.P. is supported by the F.R.S.-FNRS, Belgium.

G.H. was funded by a Rubicon grant from the Dutch Research Council (NWO).

A.K. is supported by Canada First Research Excellence Fund through the TransMedTech Institute, Canadian Open Neuroscience Platform (CONP) and International Society for Magnetic Resonance in Medicine (ISMRM).

## Author contributions

A.K., G.H. and K.W. prepared the original manuscript; A.K., G.H. and K.W. developed the initial draft of the standard and managed community contributions. A.K. merged the extension proposal to the main BIDS specification. G.H. and K.W. supervised the project. A.K., S.A, T.A., M.B., F.F., A.K., A.L., C.M., M.M., C.P., T.S., N.S., K.W. and G.H. contributed to meetings and drafts outlining the extension proposal. A.K., S.A, T.A., M.B., F.F., A.K., A.L., C.M., M.M., C.P., T.S., N.S., K.W. and G.H. revised the original manuscript.

